# Patient Knowledge and Perception about Antibiotics in Community Pharmacy

**DOI:** 10.1101/2022.11.02.22281856

**Authors:** Priya Pritika Chand

## Abstract

**Introduction:** The emergence and spread of antibiotic resistance is on the rise around the world, posing a serious threat to public health in the twenty-first century. Several research conducted in various nations have found that the general public plays a pivotal role in the increase and spread of antibiotic resistance. The present study was designed to determine the patient knowledge and perception about antibiotics in community pharmacy.

**Methods:** 200 participants were recruited by convenience sampling from patients visiting the pharmacy with a prescription for antibiotics and those fulfilling the eligibility criteria for this research. A structured questionnaire was used to access the patient’s knowledge and perception regarding antibiotics. Data collected were analyzed using Microsoft excel and Epi Info Software version 7 which was used to determine predictors of low antibiotic knowledge.

**Results:** Overall, 200 questionnaires were analyzed. 70.5% of the respondents had an intermediate level of knowledge. Misconceptions that antibiotics would work on viral infections were reported. 82% of the respondents could correctly identify that misuse of antibiotics can cause antibiotic resistance. The age, educational level, and whether or not the participants’ were studying or working in medical field were found to be important predictors of antibiotic knowledge.

**Conclusion:** The findings of this study demonstrate that the public surveyed has misunderstandings and a lack of knowledge in some crucial aspects of prudent antibiotic use. Also, negative attitudes regarding rational use of antibiotics were evident. Educational interventions are required to promote rationale use of antibiotics among the general public.

## Introduction

Antibiotics have revolutionized healthcare since their introduction in 1928 [1]. Antibiotics have had a huge impact on morbidity and mortality caused by bacterial infections, rendering them indispensable in modern medicine [1]. However, their use has been supplemented by an increase in the incidence of resistance. In addition, several million people die due to resistant infections worldwide annually [2]. In 2014, The World Health Organization (WHO) reported antimicrobial resistance (AMR) as a worldwide public health threat that requires necessary collaborative action. The Western Pacific Region, which includes Fiji, has shown the highest rate of antibiotic resistance [3]. The irrational use of antibiotics is a driving factor of antibiotic resistance (ABR).

According to the World Health Organization (WHO) definition, medicines are used rationally when patients receive the appropriate drugs, for appropriate indications, in doses that meet their individual requirements, for an adequate duration, at the lowest effective cost both to them and the society, and with appropriate information. Irrational use of medicine occurs when one or more of these conditions is not met [4]. According to the World Health Organization (WHO), more than 60% of the total antibiotics are used in the community, and nearly half of them are misused [5].

A rise in irrational antibiotic use could be influenced by a variety of factors. Many studies have shown that public perception and knowledge of antibiotics are closely linked to non-adherence to antibiotic regimens and excessive antibiotic use [6,7]. Factors associated with public knowledge of antibiotics have been reported to be demographic characteristics, including gender [6,8-10], age [8,10-14], race [9,11], education level [8-12,14-17], family income [10,16], place of residence [15-16], as well as other factors, such as lack of advice regarding rational antibiotic use, given by a physician [18].

In November 2015, Fiji became the first country in the Pacific to develop and launch a national plan for AMR [19]. The then recently completed AMR situation analysis in Fiji highlighted key challenges:

- Limited awareness and a lack of national comprehensive policies for AMR;
- Lack of a national surveillance systems to monitor AMR and antimicrobial use; and
- Limited regulation and implementation of health system responses to AMR [19].

The plan included the five key strategies to combat, or minimize the impact of, AMR in Fiji:

- Improve awareness and understanding of AMR through effective communication, education and training.
- Strengthen nationally coordinated surveillance systems.
- Reduce the incidence of antimicrobial resistance events through improved infection prevention and control, sanitation and hygiene, and wellness measures.
- Optimize the use of antimicrobial medicines in human and animal health.
- Establish and ensure governance, sustainable investment and actions to combat AMR [19].

Public education is one of the key interventions proposed by the WHO to rationalize the use of medicines [20]. Moreover, the improvement in public awareness and understanding of issues related to antibiotics is the principal strategic objective of the WHO Global Action Plan on Antimicrobial Resistance [20]. Henceforth patient knowledge, perception and practices regarding the use of antibiotics influence the decision to seek health care, the use of antibiotics and ultimately the success of the treatment.

## Methods

A descriptive, cross-sectional study design was adopted for this research. The study was carried out in Life Pharmacy, Nausori, Fiji Islands from July 2021 to September 2021. Nausori is a densely populated area with people from all economic backgrounds and education levels. Hence, it was easier to gain access to a good number of participants for this research.

Eligibility of participants was determined through a set of inclusion criteria which were as follows:

- Individuals should be 18 years or more in age
- Should be able to communicate and write in English
- Should present with a valid prescription for antibiotics
- Individuals should be residents of Fiji Islands

This study also included participants who came to the pharmacy to collect prescribed antibiotics on behalf of their family members or relatives.

The participants for this study were selected via convenience sampling. The individuals were selected based on their eligibility and willingness to participate in this research. Consenting participants completed the questionnaire which was designed to achieve the study objectives.

### Data Collection

Once approval had been received by the Fiji human health research ethics committee and the Life Pharmacy Nausori, the study was commenced. Strict covid-safe measures were followed during the data collection process such as:

- Maintaining 2 metre distance
- Temperature checks
- Hand sanitization
- Compulsory for all participants to wear masks
- CareFiji App installed in their mobile phones

Proper covid – 19 safety measures were followed during this research.

A separate area within the pharmacy premise was designated for the research purpose in order to maintain privacy of the patients and to follow proper covid-19 safety guidelines.

Individuals who fell within the inclusion criteria were handed an information sheet by the principal investigator. The principal investigator explained the details of the study using the information sheet and the individuals were given the opportunity to ask any questions that they had regarding the research. The individuals who wished to participate were given a consent form to sign. A self – administered questionnaire was then handed to the participants to fill in. It was ensured that all questions were properly answered and if there was any doubt from the patients, it was ensured that they were clarified by the researcher.

The questionnaire was divided into three main domains. The first domain consisted of the demographic details of the participants, for instance, the age, gender, economical status. The second domain consisted of questions relating to the patient’s knowledge regarding antibiotics. The participants were to write either true or false as answers to these questions. The third domain consisted of statements relating to patients’ perception about antibiotics. This was accessed using a three point scale ranging from ‘Agree’, ‘Disagree’, or ‘Unsure’.

Once the participants had completed the questionnaires, it was collected by the researcher and quickly kept in a locked cupboard. The questionnaires were coded and no personal information was collected from the participants. A log register was also maintained by researcher in order to de-identify participants. The log register, containing the patient name and the unique code present on each questionnaire, was kept separate from the data and was only accessible to the researcher. The filled questionnaires, consent forms and log register were securely kept with the researcher in a locked drawer and were used for the purpose of this research only.

### Data Management and Analysis

All data collected were coded and stored in a computer which was password protected and only accessible by the researcher. Microsoft excel and Epi Info Software version 7 were used to analyse the data to percentages, graphs and chi - square in order to summarize and present the information collected.

The knowledge assessment test was evaluated by scoring each correct response as one (1) while an incorrect response was scored zero (O). Thus, there were 0 to 14 possible scores. The percentage distribution of the antibiotics knowledge scores were determined and then used to classify respondents’ knowledge levels based on percentiles. Those whose scores were ≤ 25th percentiles were considered to have low antibiotic knowledge while those whose scores were ≥ 75th percentiles were considered to have high antibiotics knowledge. Respondents whose knowledge scores were greater than 25th percentile but less than 75th percentile were regarded to have an intermediate knowledge level.

### Ethical Considerations

The research was only conducted once appropriate approval had been received from the Fiji National Health Research ethics and review committee, Life Pharmacy Nausori and the participants.

All data collected were used for the purpose of this study only and kept securely with the researcher.

Strict covid-19 measures were adhered to while caring out the research. These measures included: wearing face mask, hand sanitization, checking temperature of participants, maintaining 2 metres distance and having CareFiji app installed in their mobile phones. Proper covid – 19 safety measures were followed during this research as per Annex 8.

Patient names were used for reference purposes only in a separate log register and kept safely in a locked cupboard, available only to the researcher. No personal information was used in the report. The participants were given a unique identification code and these codes were used to analyse the results.

## Results

A total of 200 questionnaires were distributed to the candidates who fulfilled the inclusion criteria and were willing to participate in this research. There was a 100% response rate for this research. Majority of the respondents (42%) were people aged between 31 to 40 years. Of those 200 respondents, 82 were males while 118 were females. 58% of the respondents visited the pharmacy to collect the antibiotics for themselves while 42% were there to collect medications for someone else. Highest level of education for majority of the respondents was university (65%). Most participants were employed (62%) while only 11% of them studied or worked in a medical field. Approximately two-third of the respondents were taking antibiotics for first time. A few (17%) of the participants knew the name of the antibiotics (Table 1).

**Table 1:** Demographic characteristics of respondents.

| **Variable** | **n (%)** |
| --- | --- |
| **Age (years)** |  |
| 18- 30 | 74 (37) |
| 31 – 40 | 84 (42) |
| 41 – 50 | 29 (14.5) |
| 51 - 60 | 13 (6.5) |
| **Gender** |  |
| Male | 82 (41) |
| Female | 118 (59) |
| **Antibiotic is for:** |  |
| Self-Use | 116 (58) |
| Someone Else | 84 (42) |
| **Education Level** |  |
| University | 130 (65) |
| Secondary | 55 (27.5) |
| Primary | 5 (2.5) |
| No Schooling | 10 (5) |
| **Employment** |  |
| Employed | 124 (62) |
| Unemployed | 76 (38) |
| **Work or study in medical field** |  |
| Yes | 22 (11) |
| No | 178 (89) |
| **Taking antibiotics for first time or repeats** |  |
| First time | 152 (76) |
| Repeat | 48 (24) |
| **Know which antibiotic has been prescribed** |  |
| Yes | 34 (17) |
| No | 166 (83) |

N (Total number of participants) = 200

The antibiotic knowledge assessment test revealed that 11.5% of the respondents had low knowledge level and scored between 0 – 3 points while 70.5% (scored between 4 – 10 points) and 36% (scored between 11 – 14 points) of respondents had intermediate and high knowledge levels respectively (Table 2). The modal knowledge assessment score was 1 (Figure 1).

**Table 2:** Level of Knowledge.

| Level of Knowledge | Total Score | n (%) |
| --- | --- | --- |
| Low | 0 – 3 | 23 (11.5) |
| Intermediate | 4 – 10 | 141 (70.5) |
| High | 11 – 14 | 36 (18) |

**Figure 1:**
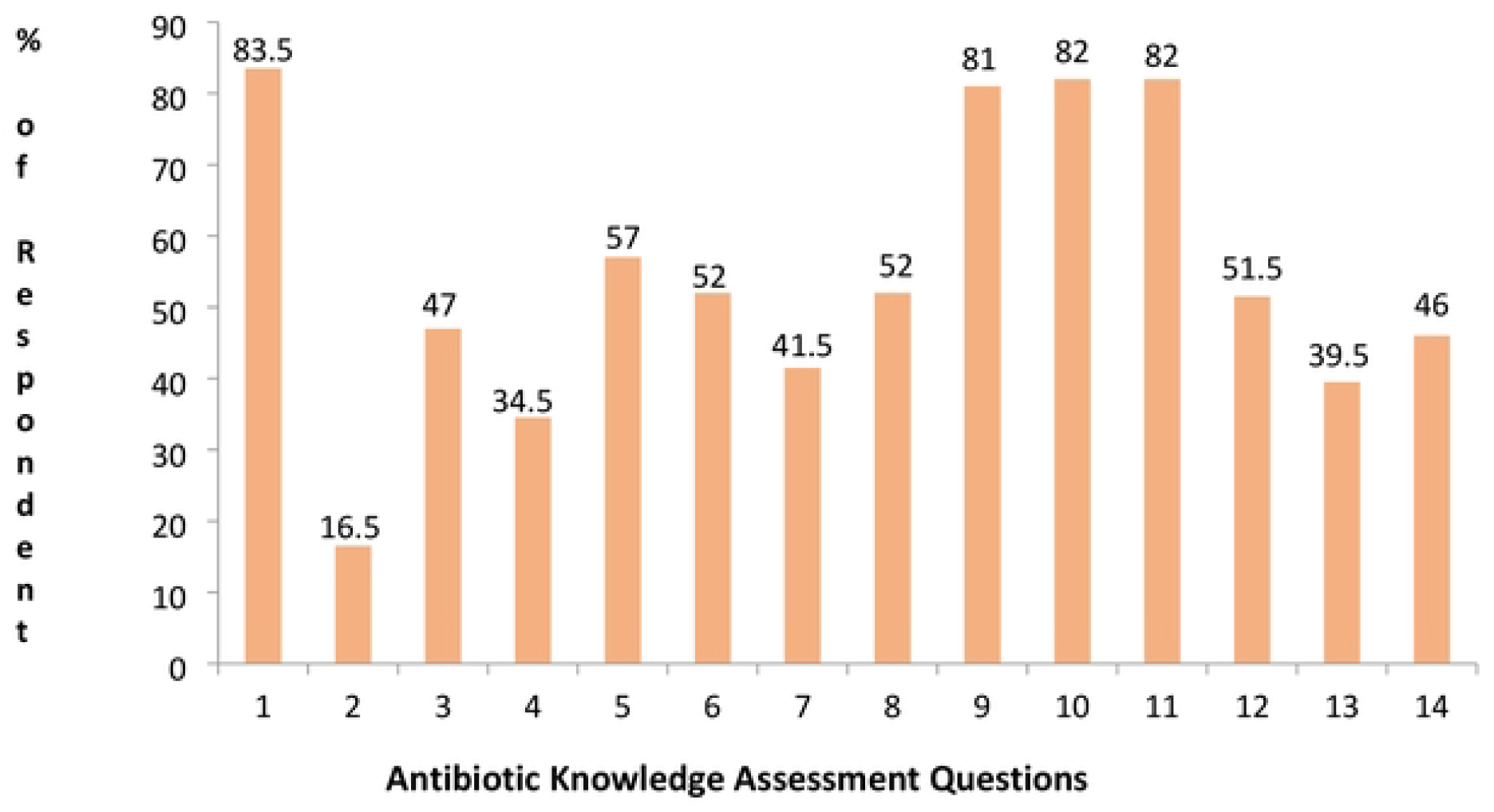
Distribution of correct knowledge responses.

Respondents in this study had a high knowledge in use of antibiotics for treating bacterial infections (83.5% correct response), identification of amoxicillin as an antibiotic (81% correct response), misuse of antibiotics can cause antibiotic resistance (82% correct response), and for some antibiotics, combination with alcohol can be dangerous (82% correct response). 83.5% of the respondents wrongly reported that antibiotics can be used to treat viral infections (Table 3).

**Table 3:** Antibiotic knowledge assessment questions and percentage of respondents with correct response.

| Knowledge assessment question | Correct Response | n (Correct Response) | % Correct Response |
| --- | --- | --- | --- |
| **Antibiotic Indications/Uses** |  |  |  |
| Antibiotics are drugs that can kill bacteria | True | 167 | 83.5 |
| Antibiotics can be used to treat viral infections | False | 33 | 16.5 |
| All infections can be cured by antibiotics | False | 94 | 47 |
| Antibiotics are used to relieve body pains | False | 69 | 34.5 |
| Antibiotics can kill the beneficial/good bacteria present on our skin and in our intestine and stomach | True | 114 | 57 |
| **Antibiotic identification** |  |  |  |
| Paracetamol is an antibiotic | False | 104 | 52 |
| Prednisolone is an antibiotic | False | 83 | 41.5 |
| Aspirin is a new generation antibiotic | False | 104 | 52 |
| Amoxicillin is an antibiotic | True | 162 | 81 |
| **Dangers associated with antibiotics** |  |  |  |
| Misuse of antibiotics can cause antibiotic resistance | True | 164 | 82 |
| For some antibiotics combination with alcohol can be dangerous | True | 164 | 82 |
| All antibiotics do not have side effects | False | 103 | 51.5 |
| **Antibiotics administration** |  |  |  |
| Antibiotics may be effective even if you did not complete treatment | False | 79 | 39.5 |
| I can reduce the dose of the antibiotic by myself if I start feeling well | False | 92 | 46 |

Low knowledge level was prevalent among respondents aged between 41 – 50 (51.7%) and 51 - 60 (61.5%). There was significant difference in the knowledge level of the two age groups with a p – value of 0.009 and 0.0005 respectively. The educational level of respondents was found to be another demographic predictor (*p* < 0.01) of low antibiotic knowledge in the study population. Based on the results obtained, respondents with no schooling or education were 6 times more likely to have low antibiotic knowledge than those with primary education (Table 4).

**Table 4:**
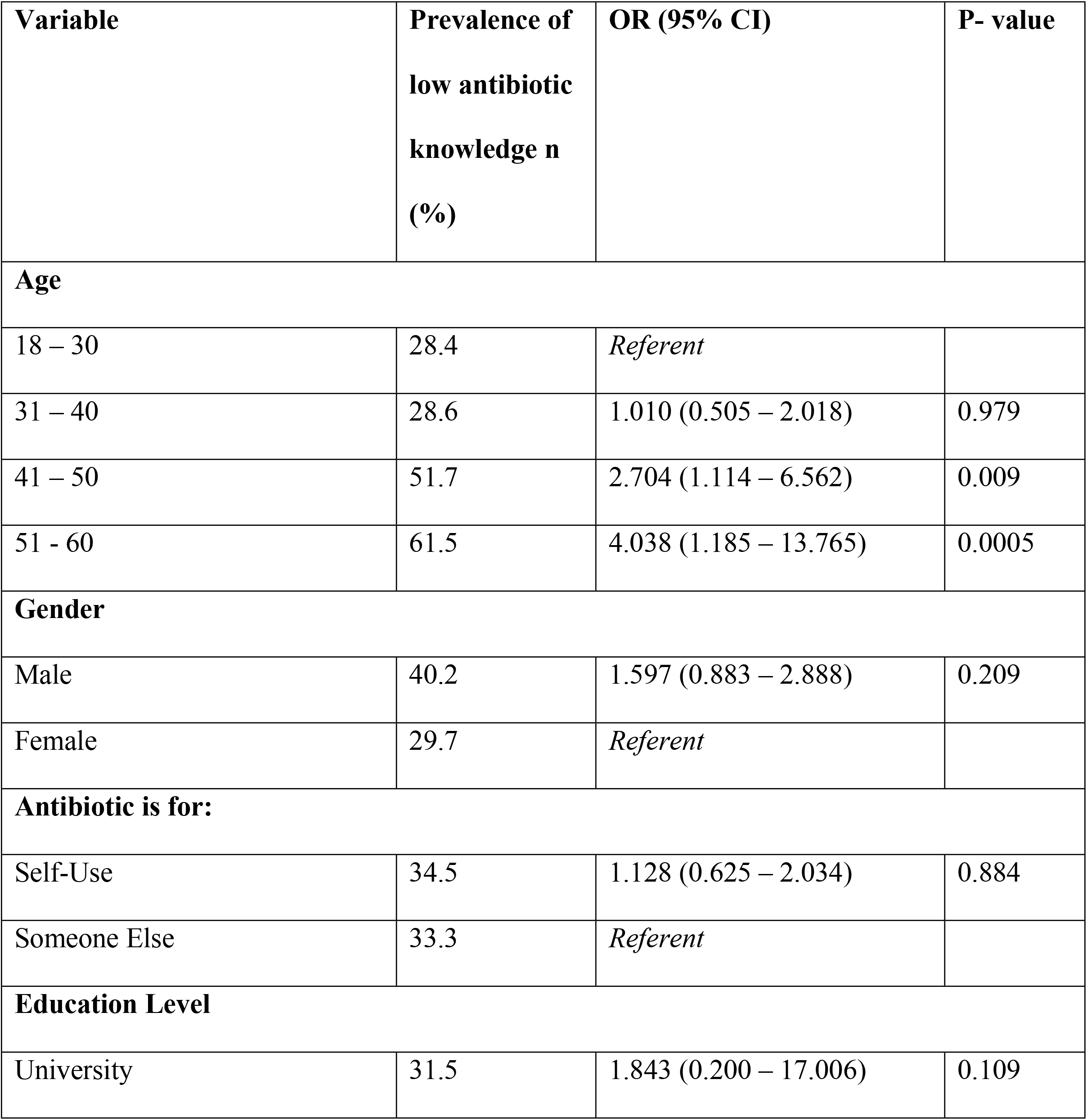
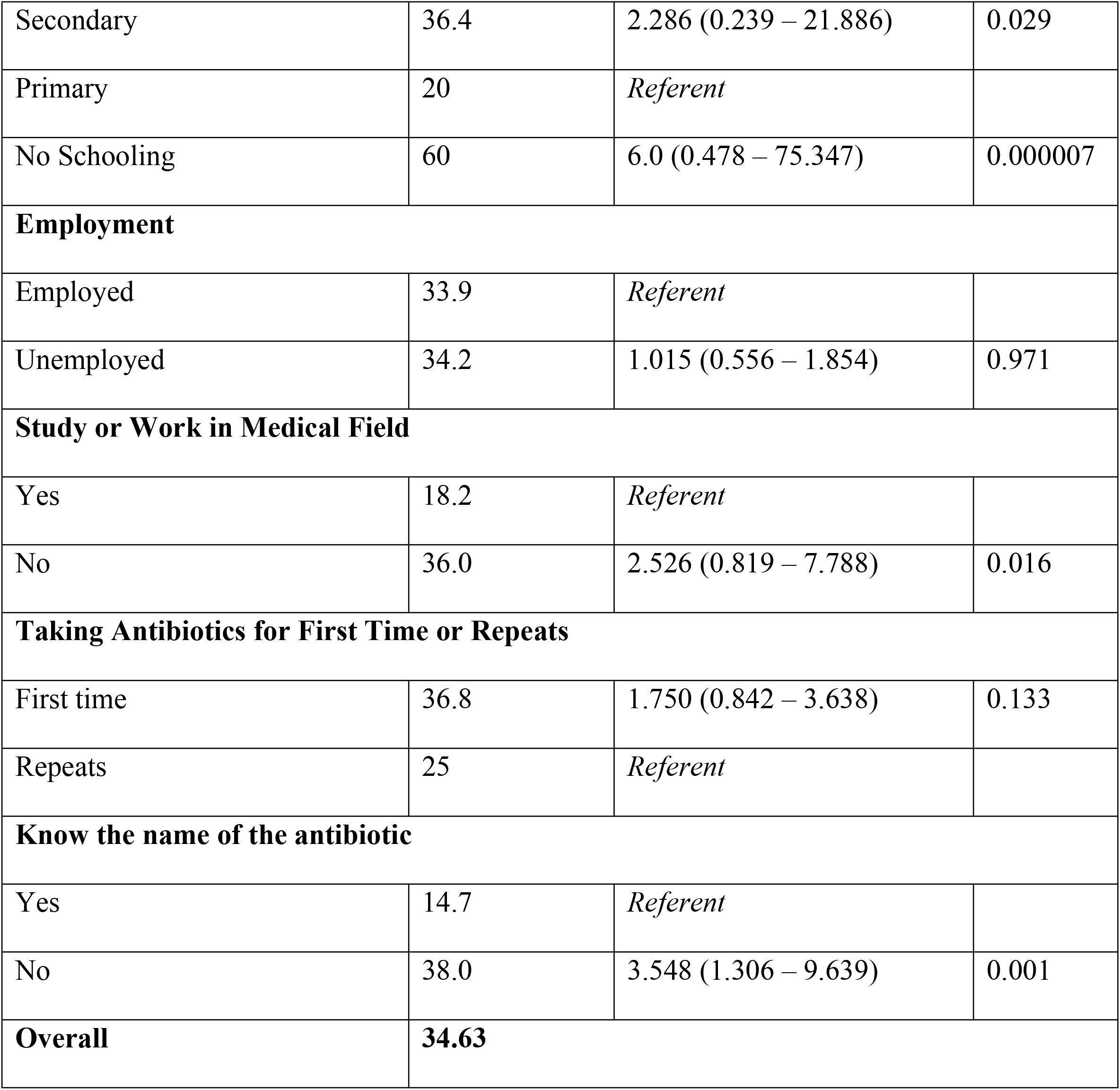
Comparison of the respondents with low antibiotic knowledge to demographic characteristics.

| Variable | Prevalence of low antibiotic knowledge n (%) | OR (95% CI) | P- value |
| --- | --- | --- | --- |
| **Age** |  |  |  |
| 18 – 30 | 28.4 | *Referent* |  |
| 31 – 40 | 28.6 | 1.010 (0.505 – 2.018) | 0.979 |
| 41 – 50 | 51.7 | 2.704 (1.114 – 6.562) | 0.009 |
| 51 - 60 | 61.5 | 4.038 (1.185 – 13.765) | 0.0005 |
| **Gender** |  |  |  |
| Male | 40.2 | 1.597 (0.883 – 2.888) | 0.209 |
| Female | 29.7 | *Referent* |  |
| **Antibiotic is for:** |  |  |  |
| Self-Use | 34.5 | 1.128 (0.625 – 2.034) | 0.884 |
| Someone Else | 33.3 | *Referent* |  |
| **Education Level** |  |  |  |
| University | 31.5 | 1.843 (0.200 – 17.006) | 0.109 |
| Secondary | 36.4 | 2.286 (0.239 – 21.886) | 0.029 |
| Primary | 20 | *Referent* |  |
| No Schooling | 60 | 6.0 (0.478 – 75.347) | 0.000007 |
| **Employment** |  |  |  |
| Employed | 33.9 | *Referent* |  |
| Unemployed | 34.2 | 1.015 (0.556 – 1.854) | 0.971 |
| **Study or Work in Medical Field** |  |  |  |
| Yes | 18.2 | *Referent* |  |
| No | 36.0 | 2.526 (0.819 – 7.788) | 0.016 |
| **Taking Antibiotics for First Time or Repeats** |  |  |  |
| First time | 36.8 | 1.750 (0.842 – 3.638) | 0.133 |
| Repeats | 25 | *Referent* |  |
| **Know the name of the antibiotic** |  |  |  |
| Yes | 14.7 | *Referent* |  |
| No | 38.0 | 3.548 (1.306 – 9.639) | 0.001 |
| **Overall** | **34.63** |  |  |

Respondents were generally found to have more positive perception towards antibiotics with results of 50% and more for majority of the dimensions studied (Table 5). The most common negative perception demonstrated by the respondents were their expectation to be prescribed an antibiotic when they suffer from flu and not taking antibiotics after they start feeling better.

**Table 5:** Perception of respondents towards antibiotic use.

| **Perception Statement\*** | **Agree**<br>**n (%)** | **Disagree**<br>**n (%)** | **Unsure**<br>**n (%)** | **Demographic factors**<br>**associated with**<br>**statement**<br>**(*p* – value)** |
| --- | --- | --- | --- | --- |
| Unnecessary use of antibiotics can make them ineffective over time | **135**<br>**(67.5)** | 33 (16.5) | 32 (16) | Age (0.006)<br>Work or study in medical field (0.034) |
| If I suffer from flu, I expect my doctor to prescribe antibiotics for me | 132<br>(66) | **49 (24.5)** | 19 (9.5) | Education (<0.001)<br>Work or study in medical field (<0.001) |
| I normally stop taking antibiotics when I start feeling better | 127<br>(63.5) | **63 (31.5)** | 10 (5) | Age (0.048)<br>Education (<0.001)<br>Work or study in medical field (<0.001) |
| If any of my family member is sick, I will give him my antibiotics if he/she has same illness as mine | 67<br>(33.5) | **120 (60)** | 13 (6.5) | None |
| I normally keep leftover antibiotics at home in case of emergency | 97<br>(48.5) | **92 (46)** | 11 (5.5) | Gender (0.009)<br>Work or study in medical field (0.005) |
| I can use the leftover antibiotics when I am suffering from a similar condition in the future | 85<br>(42.5) | **100 (50)** | 15 (7.5) | Gender (0.011)<br>Education (<0.001)<br>Work or study in medical field (0.005) |
| I will take antibiotics according to the instruction written on the label | **178 (89)** | 16 (8) | 6 (3) | None |
| Normally I check the expiry date of the antibiotic before using it. | **151 (75.5)** | 42 (21) | 7 (3.5) | Age (0.011) |

**\*Respondents with positive attitude indicated in bold**

However, respondents demonstrated positive attitudes in taking the antibiotics according to the instruction written on the label (89%), checking the expiry dates of the antibiotics before using it (75.5%), having an understanding that unnecessary use of antibiotics can make them ineffective over time (67.5%), and not giving their antibiotics to other family members with same illness (60%).

## Discussion

This study sought to identify the knowledge and perception of the Fijian general population regarding antibiotics issues and to ascertain if there were factors associated with these main outcomes of interest.

The findings imply that there are widespread misunderstandings about antibiotic use, which could result in an unnecessary risk of antibiotic-resistant infection. The most critical misconception was about the role of antibiotics in treating infections, with over 80% of those polled failing to recognize that antibiotics do not cure viral infections. The proportion of respondents that incorrectly thought that antibiotics are used in the treatment of viral infections (83.5%) is comparable to that obtained in a study in Malaysia (86.6%) [11]. In contrast, the proportion was reported to be 46.2% in a Kuwait study [7]. As a result, people must be educated about the differences between viral and bacterial infections, as well as advised not to take antibiotics for viral illnesses.

The respondents lacked the ability to distinguish between antibiotics and other regularly used medications, according to the findings. 81% of the respondents could correctly identify that amoxicillin is an antibiotic. However, paracetamol, prednisolone and aspirin were correctly identified as not being an antibiotic by 52%, 41.5%, and 52% of the respondents respectively. Several factors may have contributed to the public’s lack of understanding in this area: the public was more familiar with trade names than generic names, had never heard of or used these medicines, rarely took note of the names of the medicines they were taking, or did not receive enough information from health-care providers.

65.5% of the respondents believed that antibiotics are used to relieve body pains, 60.5% stated that antibiotics may be effective even if you did not complete treatment, while 54% declared that they can reduce the dose of the antibiotics by themselves if they start feeling well. The significant correlations found between the above mentioned knowledge statements suggest that the knowledge gap isn’t entirely random. Respondents may have mistaken antibiotics as equivalent to painkillers, prompting them to believe that discontinuing antibiotics is fine once symptoms improve, much as they would with painkillers.

In the knowledge assessment category, about two in three respondents had intermediate antibiotic knowledge. The age, educational level, and whether the participants’ were studying or working in medical field were found to be predictors of low antibiotic knowledge (Table 4). A study done in Nigeria [21] also found education level of respondents to be a predictor of low antibiotic knowledge level.

Furthermore, only 24.5% of the respondents correctly answered that they would not expect their doctor to prescribe antibiotics if they suffer from flu. Further to this, only 31.5% of the respondents said that they would not stop taking their course of antibiotics when they start feeling better, leaving an alarmingly large number (68.5%) who may stop their course of antibiotics. This high rate of participants stopping their antibiotic treatment could be a major factor leading to rapid increase in resistant bacterial infections in Fiji.

In this study, respondents demonstrated a high positive perception (75.5%) towards checking of the expiry dates of the antibiotics before using them. However, the proportion of people checking expiry dates of medication before using them is lower as compared to similar studies done in other countries: Malaysia (93%) [9], Malaysia (92.2%) [11], Lebanon (83.2%) [16], and Nigeria (93.3%) [21]. This positive perception could be a result of increased awareness by the Antimicrobial Resistance Committee, Fiji.

Unfortunately, the harm caused by antibiotic misuse extends beyond the harm caused by the medicine itself. The damage includes eliminating beneficial bacteria in our bodies (for example, in the GI tract), disrupting the immune system’s regular functioning, and a slew of other side effects. Antibiotic misuse also has a number of negative consequences, including an increase in physician visits, absenteeism, the duration of disease and suffering, and rising medical and therapeutic expenditures.

## Conclusion

70.5% of the respondents had an intermediate level of knowledge about antibiotics. 83.5% of the respondents were able to identify that antibiotics are indicated for the treatment of bacterial infections, whereas the same percentage of respondents incorrectly thought that antibiotics are also used to treat viral infections. The public surveyed had deficits in some crucial aspects of prudent antibiotic use, as well as negative attitudes about rational use of antibiotics. The findings of this study indicated areas of misconception and certain demographics in Fiji that should be targeted for educational interventions regarding the appropriate use of antibiotics.

## Data Availability

The data is included in the article.

## Acknowledgements

I thank the respondents who have taken time to participate in this research. I would also like to thank my research supervisor, Miss Ranita Kumar, for her continuous guidance throughout the research. A special thanks to Ms Numa Vera for her guidance and assistance for this research.

## Supporting Documents

S1 Figure 1. Distribution of correct knowledge responses.

S1 Appendix. Information Sheet.

S2 Appendix. Consent Form (Participants).

S3 Appendix. Approval Letter – Fiji Human Health Research and Ethics Review Committee.

S4 Appendix. Consent Form (Life Pharmacy Nausori).

S5 Appendix. Questionnaire.

S6 Appendix. Log Register.

## References

1. Almeida Santimano NM, Foxcroft DR. Poor health knowledge and behaviour is a risk for the spread of antibiotic resistance: survey of higher secondary school students in Goa, India. Perspect Public Health. 2017;137(2):109–113.

2. O’NEILL, J. Tackling Drug-Resistant Infections Globally: Final Report and Recommendations. 2016. Available online: https://amr-review.org/sites/default/files/160525_Final%20paper_with%20cover.pdf

3. The Government of Fiji. Fiji National Antimicrobial Resistance Action Plan. Suva: Ministry of health and medical services; 2015 p. 7–8. http://extwprlegs1.fao.org/docs/pdf/fij169634.pdf

4. WHO. The World Medicines Situation 2011. Medicines prices, availability and affordability; World Health Organization: Geneva, Switzerland. 2011;32.

5. Tangcharoensathien, V.; Chanvatik, S.; Sommanustweechai, A. Complex determinants of inappropriate use of antibiotics. Bull. World Health Organ. 2018, 96, 141–144.

6. Chan YH, Fan MM, Fok CM, Lok ZL, et al. Antibiotics nonadherence and knowledge in a community with the world’s leading prevalence of antibiotics resistance: Implications for public health intervention. Amer. J Infect Control. 2012 Mar 1; 40(2):113–7.

7. Awad AI, Aboud EA. Knowledge, attitude and practice towards antibiotic use among the public in Kuwait. PLoS One. 2015; 10(2):1–15.

8. Hoffmann K, Ristl R, Heschl L, Stelzer D, Maier M. Antibiotics and their effects: What do patients know and what is their source of information? Eur J Public Health. 2014; 24(3):502–7.

9. Ka Keat L, Chew Charn T. A cross sectional study of public knowledge and attitude towards antibiotics in Putrajaya, Malaysia. South Med Rev [Internet]. 2012; 5(2):26–33. Available from: http://www.fmhs.auckland.ac.nz/sop/smr/issues.aspx%5Cn http://ovidsp.ovid.com/ovidweb.cgi?T=JS&PAGE=reference&D=emed11&NEWS=N&AN=2013125286

10. Barah F, Gonçalves V. Antibiotic use and knowledge in the community in Kalamoon, Syrian Arab Republic: A cross-sectional study. East Mediterr Heal J. 2010; 16(5):516–21.

11. Oh AL, Hassali MA, et al. Public knowledge and attitudes towards antibiotic usage: A cross-sectional study among the general public in the state of Penang, Malaysia. J Infect Dev Ctries. 2011; 5(5):338–47.

12. Jose J, Jimmy B, Mohammed Saif AlSabahi AG, Al Sabei GA. A study assessing public knowledge, belief and behavior of antibiotic use in an Omani population. Oman Med J. 2013; 28(5):324–30.

13. Napolitano F, Izzo MT, Di Giuseppe G, Angelillo IF. Public knowledge, attitudes, and experience regarding the use of antibiotics in Italy. PLoS One. 2013; 8(12):1–6.

14. Kim SS, Moon S, Kim EJ. Public Knowledge and Attitudes Regarding Antibiotic Use in South Korea. J Korean Acad Nurs. 2011; 41(6):742–749.

15. Godycki-Cwirko M, Cals JWL, Francis N, et al. Public beliefs on antibiotics and symptoms of respiratory tract infections among rural and urban population in Poland: A questionnaire study. PLoS One. 2014; 9(10):1–6.

16. Mouhieddine TH, Olleik Z, Itani MM, et al. Assessing the Lebanese population for their knowledge, attitudes and practices of antibiotic usage. J Infect Public Health [Internet]. 2015; 8(1):20–31. Available from: http://dx.doi.org/10.1016/j.jiph.2014.07.010

17. Alzoubi K, Al-Azzam S, Alhusban A, et al. An audit on the knowledge, beliefs and attitudes about the uses and side-effects of antibiotics among outpatients attending 2 teaching hospitals in Jordan. East Mediterr Heal J. 2013; 19(5):478–84.

18. Fernandes M, Leite A, Basto M, et al. Non-adherence to antibiotic therapy in patients visiting community pharmacies. Int J Clin Pharm [Internet]. 2014 Oct 8 [cited 2021 Mar 18]; 36(1):86–91.

19. Fiji Launches National Antimicrobial Resistance Action Plan [Internet]. [cited 2021 Mar 12]. Available from: https://www.who.int/westernpacific/about/how-we-work/pacific-support/news/detail/18-11-2015-fiji-launches-national-antimicrobial-resistance-action-plan

20. World Health Organization. The Pursuit of Responsible Use of Medicines: Sharing and Learning from Country Experiences The benefits of responsible use of medicines: Setting policies for better and cost-effective health care [Internet]. 2012 [cited 2021 Mar 12]. Available from: https://www.who.int

21. Auta A, Banwat SB, et al. Antibiotic use in some Nigerian communities: Knowledge and attitudes of consumers. Trop J Pharm Res. 2013; 12(6):1087–92.

22. Atif M, Asghar S, et al. What drives inappropriate use of antibiotics? A mixed methods study from Bahawalpur, Pakistan. Infect Drug Resist. 2019; 12:687–99.

23. Khan FU, Khan FU, Hayat K, et al. Knowledge, attitude and practices among consumers toward antibiotics use and antibiotic resistance in Swat, Khyber-Pakhtunkhwa, Pakistan. Expert Rev Anti Infect Ther [Internet]. 2020;18(9):937–46. Available from: https://doi.org/10.1080/14787210.2020.1769477

24. Horvat OJ, Tomas AD, Kusturica MMP, et al. Is the level of knowledge a predictor of rational antibiotic use in Serbia? PLoS One. 2017;12(7):1–13.

25. Raupach-Rosin H, Rübsamen N, Schütte G, et al. Knowledge on antibiotic use, self-reported adherence to antibiotic intake, and knowledge on multi-drug resistant pathogens -results of a population-based survey in lower saxony, Germany. Front Microbiol. 2019; 10(APR):1–8.

26. OMS. Antibiotic Resistance: Multi-Country Public Awareness Survey [Internet]. WHO Press. 2015. Available from: http://apps.who.int/iris/bitstream/10665/194460/1/9789241509817_eng.pdf?ua=1

27. Farley E, Van den Bergh D, Coetzee R, Stewart A, Boyles T. Knowledge, attitudes and perceptions of antibiotic use and resistance among patients in South Africa: A cross-sectional study. South African J Infect Dis. 2019; 34(1).

28. Irawati L, Alrasheedy AA, Hassali MA, Saleem F. Low-income community knowledge, attitudes and perceptions regarding antibiotics and antibiotic resistance in Jelutong District, Penang, Malaysia: A qualitative study. BMC Public Health. 2019; 19(1):1–15.

29. Maheshwari P, Praveen D, Ravichandiran V. A study on patients awareness on rational use of antibiotics and its resistance. Asian J Pharm Clin Res. 2015; 8(3):1–4.

30. Li P, Hayat K, Shi L, Lambojon K, Saeed A, et al. Knowledge, attitude, and practices of antibiotics and antibiotic resistance among chinese pharmacy customers: A multicenter survey study. Antibiotics. 2020; 9(4):1–12.

31. Gualano MR, Gili R, Scaioli G, Bert F, Siliquini R. General population’s knowledge and attitudes about antibiotics: A systematic review and meta-analysis. Pharmacoepidemiol Drug Saf. 2015; 24(1):2–10.

32. Drozd M, Drozd K, Filip R, Byś A. Knowledge, attitude and perception regarding antibiotics among polish patients. Acta Pol Pharm -Drug Res. 2015; 72(4):807–17.

33. André M, Vernby Å, Berg J, Lundborg CS. A survey of public knowledge and awareness related to antibiotic use and resistance in Sweden. J Antimicrob Chemother. 2010; 65(6):1292–6.

34. Vallin M, Polyzoi M, Marrone G, et al. Knowledge and attitudes towards antibiotic use and resistance -A latent class analysis of a Swedish population-based sample. PLoS One. 2016; 11(4):1–18.

35. Making Your Workplace Covid-19 Safe for You & Your Customers. Min of Health & Medical Services – Fiji. 2020; (April):1–4. https://www.health.gov.fj/wp-content/uploads/2020/05/Fiji-Workplace-Preparedness-Plan-Guidelines.pdf

